# Apolipoprotein M in Right Heart Failure

**DOI:** 10.64898/2025.12.17.25342523

**Authors:** Mahmoud I Elesawy, Christina F Fu, Adeesh Parvathaneni, Arick C Park, Zhen Guo, Jason Liu, Christina Christoffersen, Carla J Weinheimer, Attila Kovacs, Jessica Nigro, Ali Javaheri, Bin Q Yang, Joel D Schilling

## Abstract

**Background:** Right heart failure (RHF) leads to an elevation in central venous pressure and causes hepatic congestion. Apolipoprotein M (ApoM), a hepatocyte-derived lipocalin bound to high-density lipoprotein, transports sphingosine-1-phosphate (S1P), which maintains vascular integrity and modulates inflammation. Although low ApoM predicts adverse outcomes in heart failure (HF), its role in RHF is unclear. We sought to investigate the impact of RHF on circulating ApoM, its prognostic value in RHF mortality, and its functional role in the cardio-hepatic axis.

**Methods:** Patients undergoing right heart catheterization were classified as normal, HF, or RHF. Serum ApoM and S1P were measured by ELISA. Survival was analyzed using Kaplan–Meier and Cox proportional hazards models. Meanwhile, ApoM Tg or WT mice were subjected to pulmonary artery banding (PAB) to induce right ventricular (RV) dysfunction or partial inferior vena cava ligation (pIVCL) to cause hepatic congestion. Cardiac and hepatic pathology were assessed by tissue imaging and molecular analyses.

**Results:** ApoM levels were lowest in RHF patients and inversely correlated with inflammatory markers. Each 0.01 μM increase in ApoM was associated with a 6% lower risk of mortality. PAB induced RV dysfunction and reduced serum ApoM in wild-type mice, while ApoM Tg mice showed less severe RV remodeling and improved hepatic congestion. In contrast, ApoM Tg mice subjected to pIVCL showed no significant improvement in liver pathology.

**Conclusion:** In patients with RHF and mice with RV dysfunction, circulating ApoM was reduced. Lower ApoM was independently associated with worse outcomes. Restoring ApoM expression primarily protects the heart and subsequently alleviates liver congestion, underscoring its distinct protective role in the heart–liver axis. Further investigation of the ApoM axis in RHF is warranted.

**Disclosures:** Research reported in this publication was supported by the National Center for Advancing Translational Sciences of the National Institutes of Health under Award Number TL1TR002344. The content is solely the responsibility of the authors and does not necessarily represent the official views of the National Institutes of Health.

**Graphical abstract:** 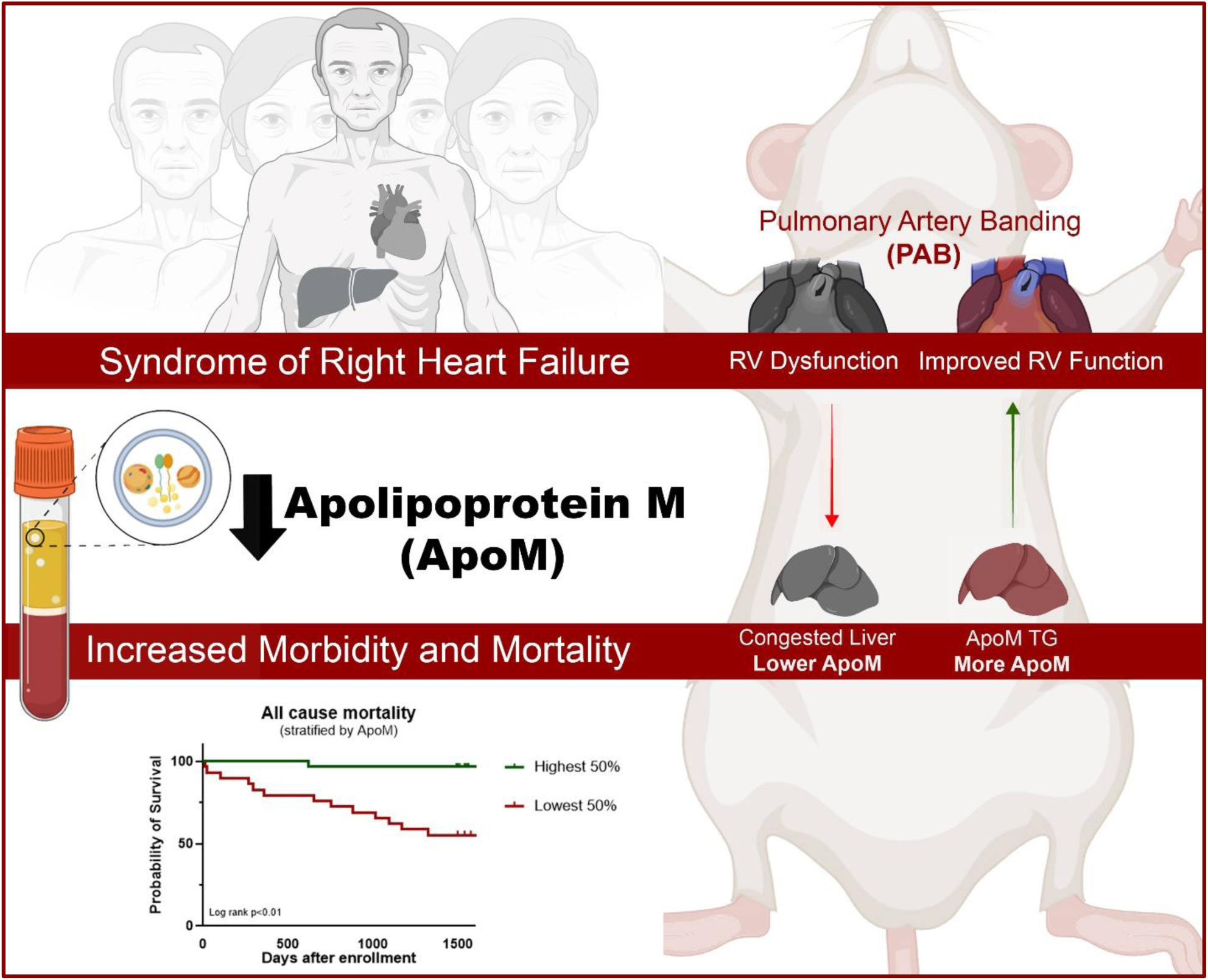

## Introduction

Heart failure (HF) is a leading cause of hospitalizations and death worldwide[1], with right heart failure (RHF) being associated with particularly poor outcomes regardless of ejection fraction[2,3]. RHF can arise due to multiple causes, including biventricular or right ventricular (RV) ischemia, cardiomyopathy, pulmonary hypertension, constrictive pericarditis, or severe tricuspid regurgitation. RHF causes elevated central venous pressure, leading to hepatic congestion. Liver congestion disrupts hepatic function, promotes biliary metaplasia, alters hepatic macrophages, and triggers fibrosis, a condition referred to as cardiogenic liver disease (CLD)[4–6]. Inflammatory changes in CLD have been shown to predict mortality in patients with HF[7].

Apolipoprotein M (ApoM) is a lipocalin predominantly produced by hepatocytes that is present on a subset of high-density lipoprotein (HDL) particles and acts as a major carrier of sphingosine-1-phosphate (S1P)[8,9]. S1P is a sphingolipid involved in cardiac and vascular signaling and in immune cell trafficking through several G-protein–coupled receptors[10]. The ApoM–S1P axis exerts anti-atherosclerotic, endothelial-stabilizing, and anti-inflammatory effects[11–14].

Reduced ApoM levels have been associated with worse outcomes in patients with HF[15]. Several cytokines have been shown to reduce ApoM expression in hepatocytes, suggesting a link between inflammation and decreased ApoM production [16,17]. However, the specific impact of RHF on ApoM expression has not been explored. In this study, we aimed to investigate the effect of RHF on circulating ApoM levels, evaluate its utility as a predictor of mortality in patients with RHF, and explore the functional impact of ApoM on the cardio-hepatic axis.

## METHODS

### Human cohort

We prospectively enrolled adult patients undergoing right heart catheterization (RHC). We excluded those with a history of transplant, infectious/inflammatory diseases, or malignancy (Fig. 1A). Patients were classified into control, HF, or RHF cohorts based on established criteria *a priori* [7]. RHF patients must have right atrial pressure (RAP) ≥ 10 mmHg, RAP/pulmonary capillary wedge pressure (PCWP) ratio > 0.55, and tricuspid annular plane excursion (TAPSE) < 1.9 cm(Fig. 1B–E). All patients were followed longitudinally for at least 3 years after RHC, and the last date of censor was November 25, 2025. The primary outcome was all-cause mortality, censoring patients who underwent LVAD implantation or heart transplantation, which were considered successful outcomes. The secondary outcome was event-free survival, defined as survival without LVAD implantation or heart transplantation. This study was approved by the Washington University Institutional Review Board (No. 201903133).

**Figure 1.**
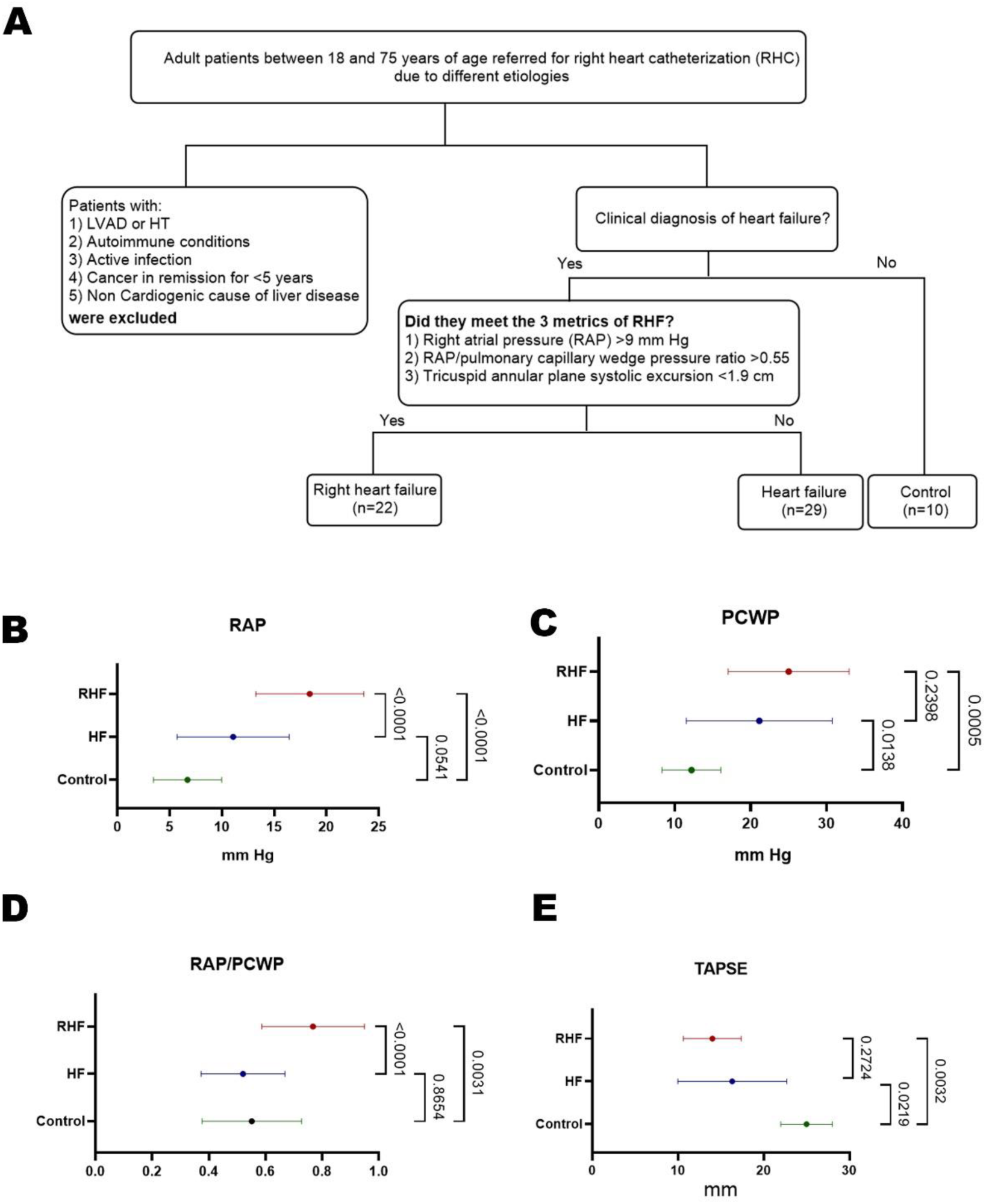
Study Design and Hemodynamic Characterization of the Patients Cohort. (A) Flowchart showing patient selection criteria for patients. (B) right atrial pressure (RAP), (C) pulmonary capillary wedge pressure (PCWP), (D) RAP/PCWP ratio, and (E) tricuspid annular plane systolic excursion (TAPSE). Data represent mean ± SD; statistical comparisons between groups were performed using one-way ANOVA. P values are indicated on the right.

### Serum analysis

Serum ApoM and S1P levels were measured by ELISA, as previously described[18], and inflammatory biomarkers were quantified via Luminex multiplex assay in the Center for Human Immunology at Washington University School of Medicine, as described [7].

### Animals

Wild-type mice C57BL/6J were purchased from Jackson Laboratory. Mice with transgenic overexpression of *huApoM* on a C57BL/6J background and littermate controls were obtained from Dr. Ali Javaheri[19]. All animals were housed in a specific pathogen-free facility under a 12-hour light/dark cycle and fed *ad libitum* with standard chow and unlimited access to drinking water throughout the study. At 8-12 weeks, they underwent pulmonary artery banding (PAB), partial inferior vena cava ligation (pIVCL), or a sham procedure. All surgical procedures were performed by the Washington University in St. Louis Mouse Cardiovascular Phenotyping Core. Briefly, for PAB, the pulmonary trunk is occluded by ∼70% luminal diameter with a ligating clip [20]; during pIVCL, a 6.0 silk thread stenosis is placed on the inferior vena cava to reduce its diameter by ∼70% [21]. After 8 weeks for pIVCL and 14 weeks for PAB, mice were anesthetized with CO_2_ and subsequently cervical dislocation. Livers and hearts were perfused with PBS via the portal vein and left ventricle, respectively, then processed for histology, RNA isolation, and flow cytometry as noted. All experimental procedures were approved by the Washington University in St Louis Institutional Animal Care and Use Committee.

### Histology

The hearts and the caudate processes and right lateral lobes of livers were fixed in 10% formalin overnight, then placed in 70% EtOH for immunohistochemistry (IHC) or 30% sucrose for immunofluorescence (IF) staining.

### Immunohistochemistry

Mouse hearts and caudate processes were paraffinized, cross-sectioned, and stained for hematoxylin and eosin (H&E) and Picrosirius red (PSR) by the Washington University in St. Louis Tissue Analysis and Imaging Core. Images were captured with the Axio Imager 2 (Zeiss) and analyzed using ImageJ for RV area and PSR quantification.

### Western blot

Plasma ApoM western blot was performed as previously described[19]. In brief, 0.5 μL plasma was added with 8.5 μL DDW and 3 μL 4x Laemmli Sample Buffer (BioRad, #1610747) and boiled to denature the plasma. Plasma mix (10 μL) was separated by 4%–20% (w/v) sodium dodecyl sulfate polyacrylamide gel electrophoresis (SDS-PAGE, BioRad, #4561093) and transferred onto polyvinylidene fluoride (PVDF) membranes (Merck Millipore, #88518, Temecula, CA, USA). The membranes were blocked with 5% (w/v) non-fat milk at room temperature for 1h and incubated overnight with 1:1000 anti-mouse ApoM antibody (LS Bio, # LSC158166). Then the membranes were washed and further treated with HRP-conjugated secondary antibodies at room temperature for 1 h. The immune complexes were visualized by enhanced chemiluminescence methods. The band intensity was measured and analyzed with NIH ImageJ software (Bethesda, MD, USA).

### Immunofluorescence

Right lateral lobes were rinsed with PBS and embedded in OCT media, cut into 8 μm sections using a cryostat (Leica Biosystems), mounted onto glass slides (Lab Storage), and stored at -80 °C. For IF staining, sections were air-dried at RT for 15 min, rehydrated with PBS for 5 min, encircled with hydrophobic pen, and blocked with fresh blocking buffer (PBS with 1% BSA w/v and 0.3% Triton X-100 v/v) at RT for 1 hr. Primary antibodies were diluted in blocking buffer as follows (50 μL/section): 1:100 Rabbit anti-Desmin (Abcam, ab15200), Rat anti-F4/80 Biotin 1:250 (eBioscience, 13-4801-85), and incubated on sections at 4 °C in a humidified chamber overnight. The sections were then washed with PBS three times for 5 min each. Secondary antibody solution was made in blocking buffer as follows: Goat anti-Rabbit AF 488 (1:500, Invitrogen, A11008), Donkey anti-Rat AF 594 (1:500, Invitrogen, A21209). Sections were incubated in secondary antibody solution for 1 hour at room temperature in the dark. The sections were washed with PBS three times for 5 min each, then stained with fresh Hoechst dye (1:25000 in PBS, Thermo Scientific) for 5 min in the dark. Stained sections were dried and mounted with ProLong Gold Antifade Mountant (Invitrogen) and 1.5 mm glass coverslips. Confocal images were acquired using an LSM 900 laser-scanning confocal microscope (Zeiss) and analyzed with ImageJ.

### RNA isolation and quantitative real-time polymerase chain reaction

The right liver lobes were snap frozen in liquid nitrogen on the day of tissue collection and stored at -80 °C until RNA extraction. RNA was isolated with the PureLink RNA kit (Life Technologies), and the NanoDrop One (Thermo Scientific) was used to measure concentration and purity. cDNA was reverse transcribed from 0.5 μg RNA using random oligonucleotide primers and the Reverse Transcriptase kit (Applied Biosystems). Quantitative real-time polymerase chain reaction (PCR) was conducted in duplicate measurements using Power SYBR Green PCR Master Mix (Applied Biosystems) and the QuantStudio3 Real-Time PCR System (Thermo Scientific). Data were analyzed using the ΔΔCt method and normalized to *36b4* gene expression. The following primers were used (5′ to 3′):

*Acta2* forward: GTCCCAGACATCAGGGAGTAA

and reverse: TCGGATACTTCAGCGTCAGGA,

*Mmp2* forward: CAAGGATGGACTCCTGGCACAT

and reverse: TACTCGCCATCAGCGTTCCCAT,

*Ck19* forward: GGGGGTTCAGTACGCATTGG

and reverse: GAGGACGAGGTCACGAAGC,

*Ck7* forward: AGGAGATCAACCGACGCAC and

reverse: GTCTCGTGAAGGGTCTTGAGG,

*Col1a2* forward GCTCCTCTTAGGGGCCACT and

reverse: CCACGTCTCACCATTGGGG,

*Col3a1* forward CTGTAACATGGAAACTGGGGAAA and

reverse: CCATAGCTGAACTGAAAACCACC, and

*36b4* forward: ATCCCTGACGCACCGCCGTGA and

reverse: TGCATCTGCTTGGAGCCCACGTT.

### Statistical analysis

Categorical variables were compared using Fisher’s exact test, and continuous variables were analyzed with Student’s t-test. For comparisons involving more than two groups, one-way ANOVA was performed. Normality was assessed using the Shapiro–Wilk test. Kaplan–Meier curves were generated with endpoints of death or LVAD/ heart transplant.

Groups were stratified by the median values of selected variables, and differences between survival curves were assessed using the log-rank test. For multivariable analyses, a Cox proportional hazards model was applied to the composite endpoint of death, LVAD implantation, or heart transplantation, with adjustment for age, creatinine, Bilirubin, and left ventricular ejection fraction (LVEF). ApoM concentrations were multiplied by 10 before Cox regression analysis to improve numerical scaling. Pearson correlation analyses were performed to assess associations between ApoM and various cytokines and markers of end-organ injury. All statistical analyses were performed using GraphPad Prism version 9.3.0 (GraphPad Software, San Diego, CA), and all graphics were created using BioRender.com and Adobe Photoshop version 26.11.

## Results

### ApoM is reduced in patients with RHF

To assess circulating ApoM levels, we collected samples from controls and patients with HF and RHF at the time of right heart catheterization (RHC). Baseline demographics and hemodynamics for the three patient cohorts are summarized in Table 1. As expected, the RHF group showed higher RAP and greater RV chamber dilation. RV systolic dysfunction was also more significant in patients with RHF, reflected by lower TAPSE and a high RAP/PCWP ratio. Regarding liver function, bilirubin and INR levels were higher, and albumin was lower in both HF and RHF compared with controls, with no difference between HF and RHF. Aspartate aminotransferase (AST) and alanine aminotransferase (ALT) did not differ between groups.

**Table 1.** Baseline demographics of normal controls, patients with heart failure (HF) but preserved right ventricular function, and patients with right heart failure (RHF).

| **Table 1.** | **Control**<br>**(N=10)** | **HF**<br>**(N=29)** | **RHF**<br>**(N=22)** |
| --- | --- | --- | --- |
| **Demographics** | **Mean (SD)** | **Mean (SD)** | **Mean (SD)** |
| Age (years) | 49.5 (16.4) | 53.3 (7.7) | 59.7 (11.8) |
| BMI (kg/m<sup>2</sup>) | 29.1 (3.9) | 30.2 (8.8) | 29.9 (7.5) |
| Gender * | **n (%)** | **n (%)** | **n (%)** |
| Male | 3 (30) | 23 (79) | 12 (55) |
| Female | 7 (70) | 6 (21) | 10 (45) |
| Race |  |  |  |
| White | 8 (80) | 19 (66) | 12 (55) |
| Black | 2 (20) | 10 (34) | 8 (36) |
| Asian |  |  | 1 (5) |
| Cardiomyopathy etiology<br>(Nonischemic) *† | -- | 22 (76) | 14 (64) |
| Hypertension | 2 (20) | 11 (38) | 9 (41) |
| Diabetes (%) | 1 (10) | 5 (17) | 9 (40) |
| CKD *† | -- | 11 (38) | 13 (60) |
| Hyperlipidemia | 2 (20) | 8 (28) | 4 (18) |
| **Hemodynamic and Echocardiogram Data** | **Mean (SD)** | **Mean (SD)** | **Mean (SD)** |
| RAP (mmHg) *†‡ | 6.7 (3.3) | 11 (5.4) | 18 (5.2) |
| PCWP (mmHg) *†‡ | 12.2 (3.9) | 21.1 (9.6) | 25 (8) |
| RAP/PCWP ratio ‡ | 0.55 (0.2) | 0.52 (0.1) | 0.77 (0.2) |
| mPAP (mmHg) *†‡ | 24.5 (7.9) | 33.4 (11.7) | 41.4 (11) |
| Cardiac output (L/min) | 5.9 (2.0) | 4.8 (2.6) | 4.2 (1.3) |
| Cardiac index (L/min/m<sup>2</sup>) *† | 3.3 (1.3) | 2.2 (1.6) | 2.1 (0.6) |
| LV ejection fraction (%) *†‡ | 63.2 (4.4) | 23 (14.5) | 38.4 (23.3) |
| TAPSE (cm) *† | 2.5 (0.3) | 1.6 (0.5) | 1.4 (3.4) |
| RV diameter (cm) †‡ | 3.9 (0.7) | 4.13 (1) | 4.69 (0.9) |
| **Lab values** | **Mean (SD)** | **Mean (SD)** | **Mean (SD)** |
| Bilirubin (mg/dL) *† | 0.4 (0.3) | 1.4 (1.1) | 1.4 (1.1) |
| Creatinine (mg/dL) *† | 0.8 (0.2) | 1.4 (0.6) | 1.6 (0.7) |
| Hemoglobin (gm/dL) | 13.2 (1.2) | 13.1 (1.6) | 11.7 (2.3) |
| Platelet ($\times 10^9$/L) | 258.9 (58.7) | 245.5 (123) | 222.3 (107.8) |
| Sodium (mEq/L) | 140.8 (1.9) | 136.5 (4.9) | 136.1 (5.7) |
| ALT (U/L) | 22.9 (9.0) | 86.6 (236.8) | 56.3 (93.7) |
| AST (U/L) | 22.5 (9.6) | 74.6 (168.5) | 48.3 (46) |
| Albumin (g/dL) *† | 4.3 (0.2) | 3.9 (0.6) | 3.8 (0.6) |
| NT-proBNP (pg/mL) *† | 188 (58.6) | 3263 (2986) | 4608 (5748) |
| INR *† | 1.1 (0.1) | 1.6 (0.5) | 1.5 (0.3) |
| HBA1c | 6.5 (1.2) | 6.3 (1.8) | 6.9 (1.4) |
| MELD-XI*† | 9.5 (0.2) | 14.3 (4.9) | 15.7 (5.6) |

Numerical variables were compared using one-way ANOVA; categorical variables using Fisher's exact test. \*P<0.05 between normal and HF, †P<0.05 between normal and RHF, ‡P<0.05 between HF and RHF. CKD: chronic kidney disease; RAP: right atrial pressure; PCWP: pulmonary capillary wedge pressure; TAPSE: tricuspid annular plane systolic excursion; mPAP: mean Pulmonary artery pressure; MELD-XI: Model for End-stage Liver Disease excluding INR; cardiac output and index (Fick method)

We found that patients with HF had lower serum ApoM compared with controls, with those with RHF having the lowest levels relative to both HF and controls (Fig. 2A). Lower ApoM was associated with reduced overall survival and decreased LVAD/transplant-free survival on Kaplan–Meier analysis (Fig. 2B–C). Of note, S1P concentrations were unchanged across groups and showed no significant association with survival (Fig. 2D–F). HDL levels were also lower in HF and RHF compared with controls but did not differ between HF and RHF. Although HDL is associated with overall survival, it did not predict LVAD/transplant–free survival (Fig. 2G–I). In addition, LDL and total cholesterol were the lowest in RHF with no difference in triglyceride levels (Sup. Fig. 1A–C). Using Cox proportional hazards modeling adjusted for age, LVEF, creatinine, and bilirubin (Fig. 2J), each 0.01 μM increase in ApoM corresponded to a 6% reduction in risk of death. This association persisted after further adjustment for different variables.

**Figure 2.**
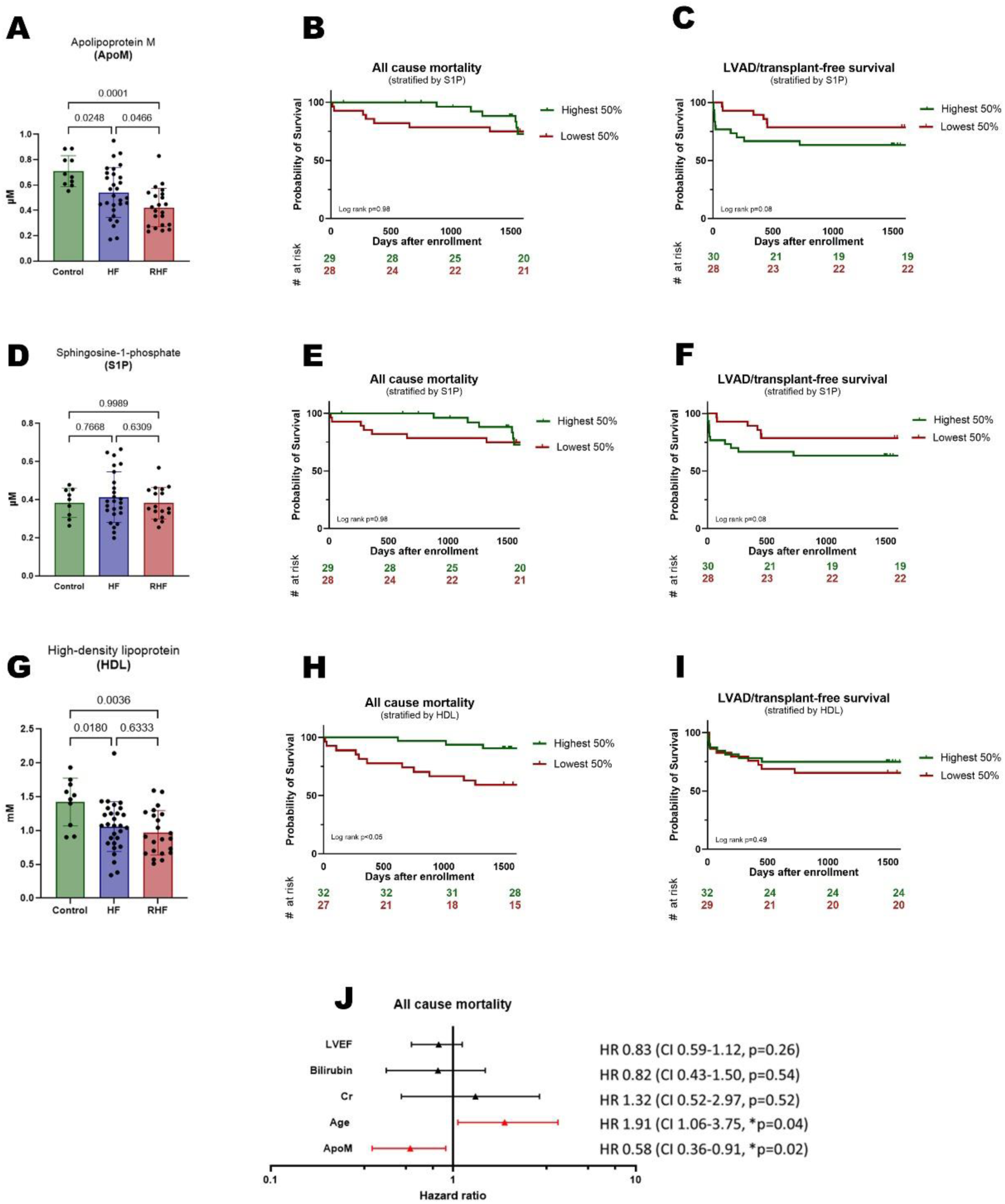
Circulating ApoM, S1P, and HDL Levels and Their Association with Survival in Heart Failure. (A) Serum ApoM levels across groups. (B) Kaplan–Meier curve for all-cause mortality stratified by ApoM. (C) Kaplan–Meier curve for LVAD/transplant-free survival stratified by ApoM. (D) Serum S1P levels across groups. (E) Kaplan–Meier curve for all-cause mortality stratified by S1P.(F) Kaplan–Meier curve for LVAD/transplant-free survival stratified by S1P. (G) Serum HDL levels across groups. (H) Kaplan–Meier curve for all-cause mortality stratified by HDL.(I) Kaplan–Meier curve for LVAD/transplant-free survival stratified by HDL. (J) Forest plot showing multivariable Cox regression analysis for predictors of all-cause mortality, including left ventricular ejection fraction (LVEF), bilirubin, creatinine (Cr), age, and ApoM. Data are presented as mean ± SD, p values by one-way ANOVA.

### ApoM levels are inversely related to markers of congestion and inflammation

Across the cohort, ApoM levels showed consistent inverse correlations with markers of venous congestion, hepatic dysfunction, and systemic inflammation. Lower ApoM was associated with higher RAP, mean pulmonary artery pressure, and PCWP (Fig. 3A–C). ApoM inversely correlated with bilirubin, MELD-XI, and ALT (Fig. 3D–F), and similarly with AST, creatinine, and NT-proBNP, while showing a positive association with albumin (Sup. Fig. 2A–D). Of particular interest, lower ApoM levels were associated with higher levels of inflammatory and macrophage activation markers, including soluble CD163, CXCL12, and CXCL10 (Fig. 3G–I) as well as VEGF-A, CCL11, MIP1β, IL-10, IL-27, and IL-2 (Sup. Fig. 3A-F).

**Figure 3.**
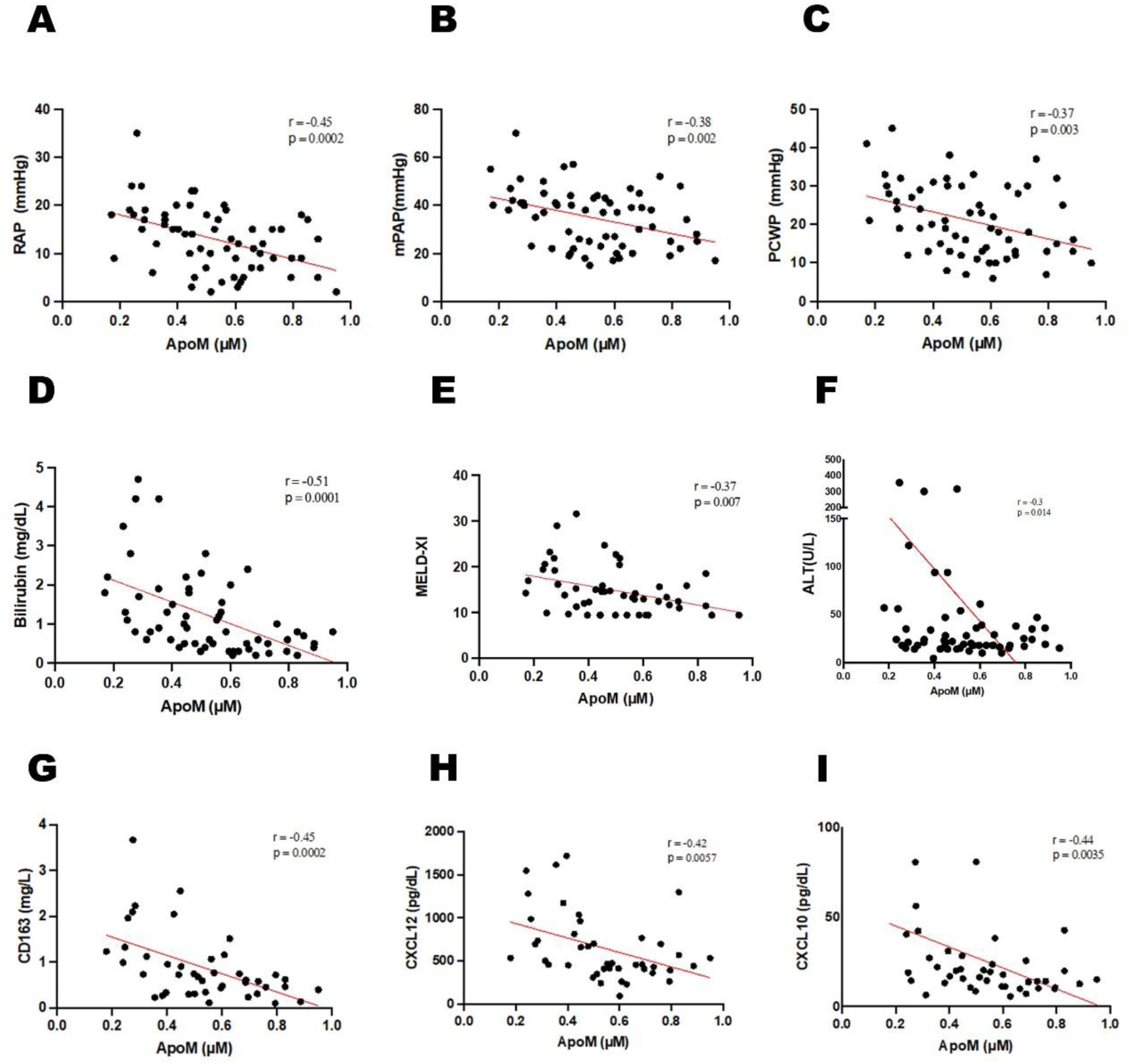
Correlations Between Serum ApoM and Hemodynamic, Liver, and Inflammatory Markers. (A) Right atrial pressure (RAP). (B) Mean pulmonary arterial pressure (mPAP). (C) Pulmonary capillary wedge pressure (PCWP). (D) Total bilirubin. (E) MELD-XI scores. (F) Alanine aminotransferase (ALT). (G) CD163. (H) CXCL12. (I) CXCL10. Pearson correlation used for analysis

### RV dysfunction is associated with liver pathology and lower ApoM

To further evaluate the relationship between RHF, ApoM, and liver pathology, we subjected mice to PAB. Echocardiography performed 14 weeks after surgery demonstrated adverse RV remodeling, including RV hypertrophy, reduced TAPSE, increased end-diastolic area, worse TR grade, and decreased RVEF compared with sham-treated animals (Fig. 4A–C). RV remodeling was further supported by histology, which demonstrated an increased relative RV area (Fig. 4D) and greater fibrosis on PSR staining (Fig. 4E). Livers from PAB mice showed dilated sinusoids and expansion of hepatic stellate cells (HSCs) (Fig. 4F–G). Several genes associated with fibrosis and ductular reaction were also upregulated in the liver after PAB (Fig. 4H). Plasma ApoM levels measured by Western blot were also significantly reduced in PAB mice compared with sham animals (Fig. 4I).

**Figure 4.**
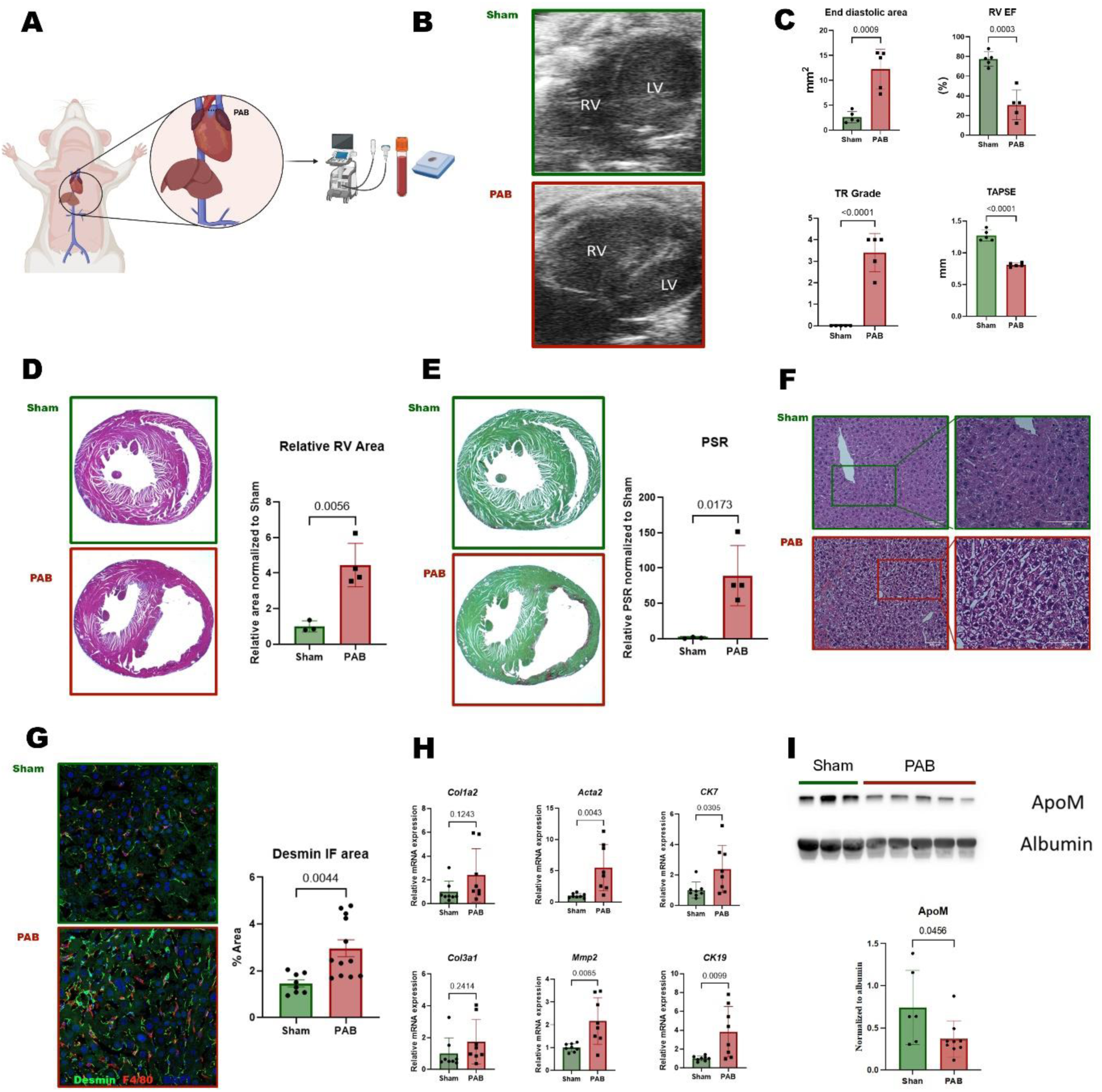
Pulmonary Artery Banding (PAB) Induces Right Ventricular Dysfunction and Hepatic Congestion. (A)Schematic representation of the PAB model. (B) Representative echocardiographic images showing right ventricular (RV) dilation in PAB versus sham mice. (C) Quantification of RV end-diastolic area, ejection fraction (EF), tricuspid regurgitation (TR) grade, and tricuspid annular plane systolic excursion (TAPSE). (D) Hematoxylin and eosin (H&E) staining showing RV dilation relative to sham. (E) Picrosirius red (PSR) staining demonstrating increased RV fibrosis in PAB mice, with quantification normalized to sham. (F) H&E staining of liver sections showing hepatic congestion in PAB mice. (G) Immunofluorescence staining for Desmin showing increased fibrotic remodeling in PAB hearts with quantification of Desmin-positive area. (H) qPCR analysis of fibrotic (*Acta2, Col3aActa2, Mmp2*), ductular reaction (*CK7, CK19*), in livers from sham and PAB mice. (I) Western blot showing reduced hepatic ApoM protein expression normalized to albumin in PAB mice. Statistical comparisons were performed using unpaired two-tailed Student’s t-test; data are presented as mean ± SD.

### ApoM improves pressure overload-induced RV dysfunction

Beyond its role as a biomarker, we sought to investigate whether ApoM levels could impact RV remodeling in response to pressure overload. Therefore, we subjected ApoM transgenic (ApoM Tg) mice and wild-type littermate controls to PAB. Echocardiography performed two months after surgery demonstrated improved RV function in ApoM Tg mice, reflected by higher TAPSE and RVEF compared with wild-type mice (Fig.5A). Histological analysis showed smaller RV relative area and reduced fibrosis via PSR staining in ApoM Tg mice (Fig. 5B–C). Additionally, ApoM Tg mice exhibited significantly less HSC expansion compared with controls (Fig. 5D), along with a downward trend of genes associated with ductular reaction and fibrosis (Sup. Fig. 4A–D).

**Figure 5.**
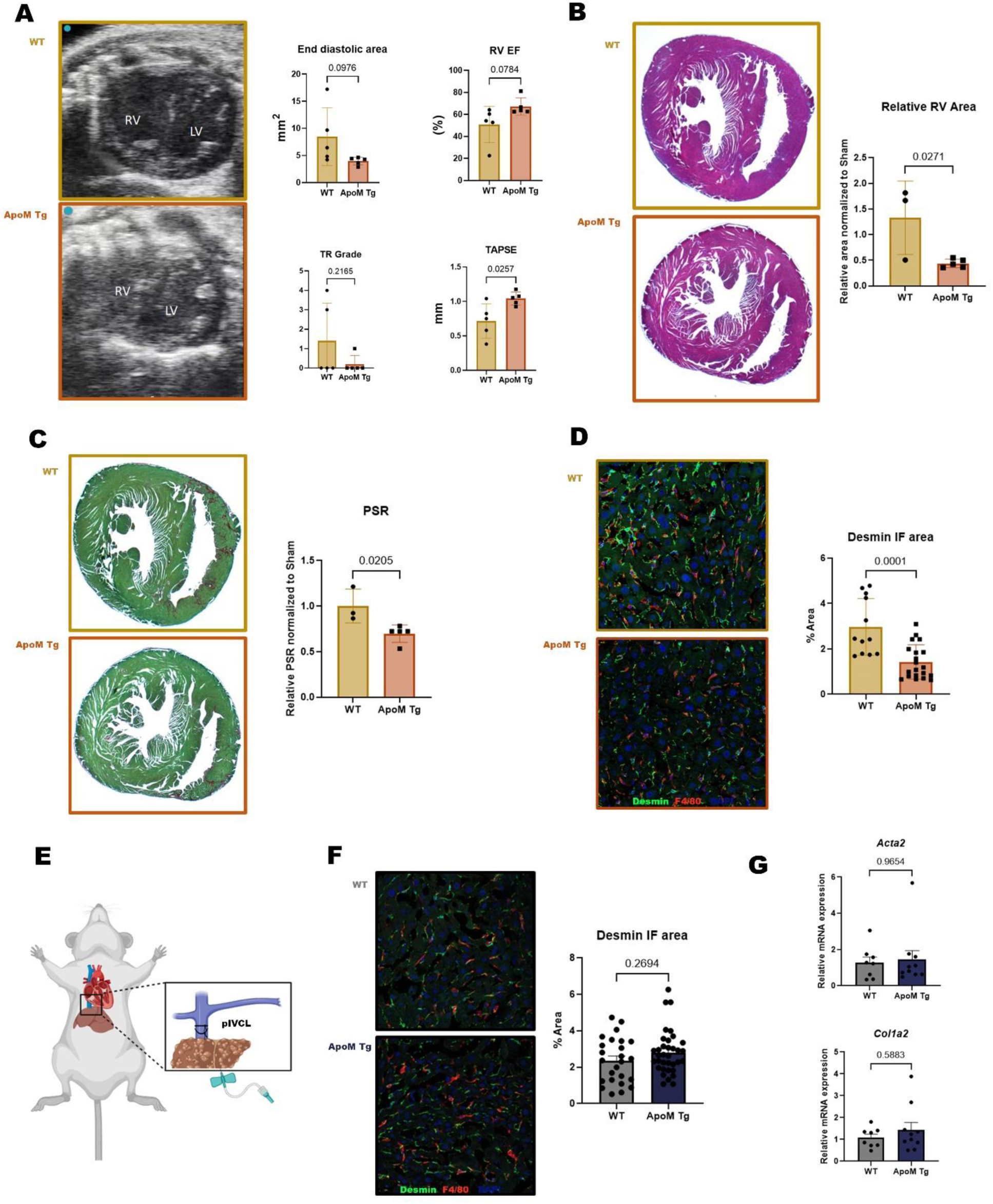
ApoM Overexpression Attenuates Right Ventricular Remodeling and Hepatic Congestion After PAB. (A) Representative echocardiographic images showing preserved right ventricular (RV) function in ApoM transgenic (Tg) mice compared with wild type (WT), with quantification of RV end-diastolic area, ejection fraction (EF), tricuspid regurgitation (TR) grade, and tricuspid annular plane systolic excursion (TAPSE). (B) Hematoxylin and eosin (H&E) staining demonstrating improved RV in ApoM TG mice with reduced RV area normalized to control. (C) Picrosirius red (PSR) staining showing decreased RV fibrosis in ApoM TG mice relative to control. (D) Immunofluorescence staining for Desmin showing reduced Desmin-positive area in ApoM TG RV tissue. (E) Schematic illustration (partial inferior vena cava ligation (pIVCL) in ApoM TG and control mice. (F) Representative Desmin immunofluorescence staining in the liver showing no significant difference between groups. (G) Hepatic mRNA expression of αSma and Col1a2 showing no difference between control and ApoM TG mice in pIVCL. Statistical comparisons were performed using unpaired two-tailed Student’s t-test; data are presented as mean ± SD.

### ApoM does not directly impact liver pathology

Based on the fact that ApoM overexpression improved markers of liver pathology after PAB, we sought to determine whether ApoM could directly mediate protective effects in the liver. Therefore, we subjected ApoM Tg and wild-type mice to a model of hepatic congestion using partial IVC ligation (pIVCL), which recapitulates features of human liver congestion and fibrosis without any cardiac pathology [6] (Fig. 5E). In contrast to PAB, ApoM Tg mice had a similar degree of HSCs activation and fibrosis compared with wild-type controls after pIVCL (Fig. 5F–G). While tissue pathology was not impacted by increased ApoM, we did observe a slight shift in liver macrophage subsets in these mice (Sup. Fig. 5A–C). We conclude that ApoM overexpression has no direct impact on liver pathology but rather improves hepatic biology via its impact on RV remodeling.

## Discussion

In this study, we investigated the relationship between ApoM and RHF. Our primary findings were that ApoM levels were reduced in patients with RHF and in mice with pressure overload–induced RV dysfunction. Enhancing ApoM expression conferred protection to the RV and secondarily improved hepatic outcomes, though it did not directly ameliorate liver function in an isolated model of hepatic congestion.

ApoM binds to HDL and carries S1P in circulation. Its biological effects are thought to be mediated by S1P-dependent mechanisms to maintain vascular integrity, lymphocyte trafficking, cell proliferation, and survival [22–24]. ApoM is predominantly produced in the liver; therefore, the reduced levels we observed with RHF may indicate that it serves as a biomarker of liver health[25]. While we found that ApoM was lowest in patients with RHF, serum S1P and the fraction of S1P bound to ApoM were unchanged. ApoM is the major carrier of S1P in circulation; however, up to 30% of S1P is carried by albumin and other lipoproteins. Thus, the distribution of S1P between HDL/ApoM and albumin could be altered in RHF, which may change S1P-receptor signaling and increase vascular inflammation [26–28].

In addition, lower ApoM is a prognosticator of adverse outcomes in HF and RHF. We found that ApoM was inversely correlated with several inflammatory markers, which may indicate an anti-inflammatory role of this lipoprotein in RHF [29]. HDL levels were associated with overall mortality, but they do not predict the need for transplant or LVAD. This suggests that the qualitative HDL dysfunction by alteration of attached lipoprotein, rather than reduced HDL levels per se, may be relevant in RHF[30–32].

While the mechanism by which ApoM levels decrease during RHF remains unknown, we did observe a negative correlation between ApoM concentration and RAP, PCWP, and mean pulmonary artery pressure; suggesting that impaired hemodynamics are associated with a reduction in ApoM. ApoM was also inversely related to several metrics of liver and kidney function, including bilirubin, MELD-XI, and creatinine, in line with the fact that ApoM may reflect chronic organ congestion in RHF. Indeed, we found that ApoM and serum inflammatory markers were inversely related. Thus, ApoM may not only be a biomarker of poor prognosis in RHF but also be related to the altered inflammatory milieu associated with hepatic congestion.[7,33,34]

Another question we addressed in this study is whether ApoM is also a contributor to pathology in RHF and/or congestive liver disease. To explore this, we subjected mice to PAB, which led to characteristic features of RHF along with evidence of early congestive hepatopathy, including sinusoidal dilatation and HSC expansion [35]. In contrast, ApoM-Tg mice, which express human ApoM at 2-3 times the level of endogenous ApoM, developed significantly less severe RV remodeling after PAB. These findings argue that reduced levels of ApoM are more than a prognostic marker in HF but could also contribute to the progression of the disease. ApoM-Tg mice also had less expansion of HSC, the principal fibrogenic cellular population in the liver[36,37]. However, the fact that ApoM overexpression did not impact liver remodeling after pIVCL indicates that the beneficial impact of this lipoprotein on liver pathology occurs primarily as a consequence of its effects on the RV.

Together, our findings highlight a unique aspect of the cardio-hepatic axis in heart failure. Specifically, the liver is the main source of HDL and ApoM. However, during the progression of RHF, ApoM levels decrease, which has the potential to exacerbate adverse RV remodeling. Delineating the molecular mechanisms behind the drop in ApoM in RHF and evaluating therapies designed to increase ApoM are important areas for future investigation.

There are several important limitations of our study, including the relatively small sample size from a single center. Additionally, although we aimed to recruit the healthiest possible controls, they were still individuals undergoing right heart catheterization for clinical reasons. The ApoM Tg mice increase the levels of ApoM throughout the life of the animal and therefore could change baseline heart biology before PAB.

In conclusion, we found that ApoM levels decrease in RHF, and this predicts adverse outcomes in HF patients. Moreover, increasing ApoM levels in a mouse model of RHF improved RV pathology, suggesting that this axis could be modulated for therapeutic benefit. Further investigation of the ApoM axis in RHF is warranted.

## Data Availability

The data that support the findings of this study are available from the corresponding author upon reasonable request.

## Supplemental Figures

**Supplemental Figure 1.**
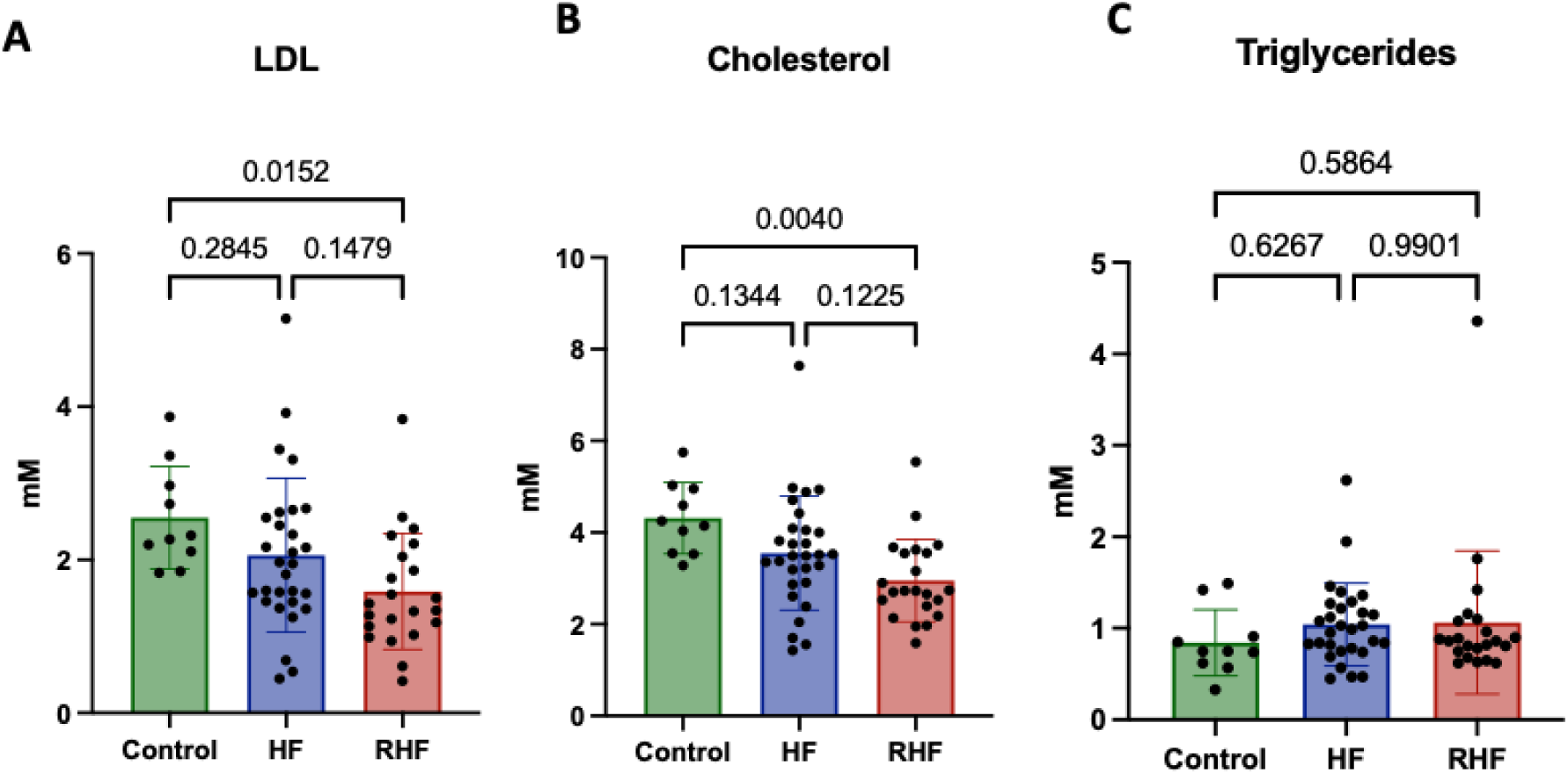
Serum lipid profile in patients with control, heart failure (HF), and right heart failure (RHF) groups. (A) LDL, (B) Total cholesterol, and (C) Triglycerides. Data are presented as mean ± SD, p values by one-way ANOVA.

**Supplemental Figure 2.**
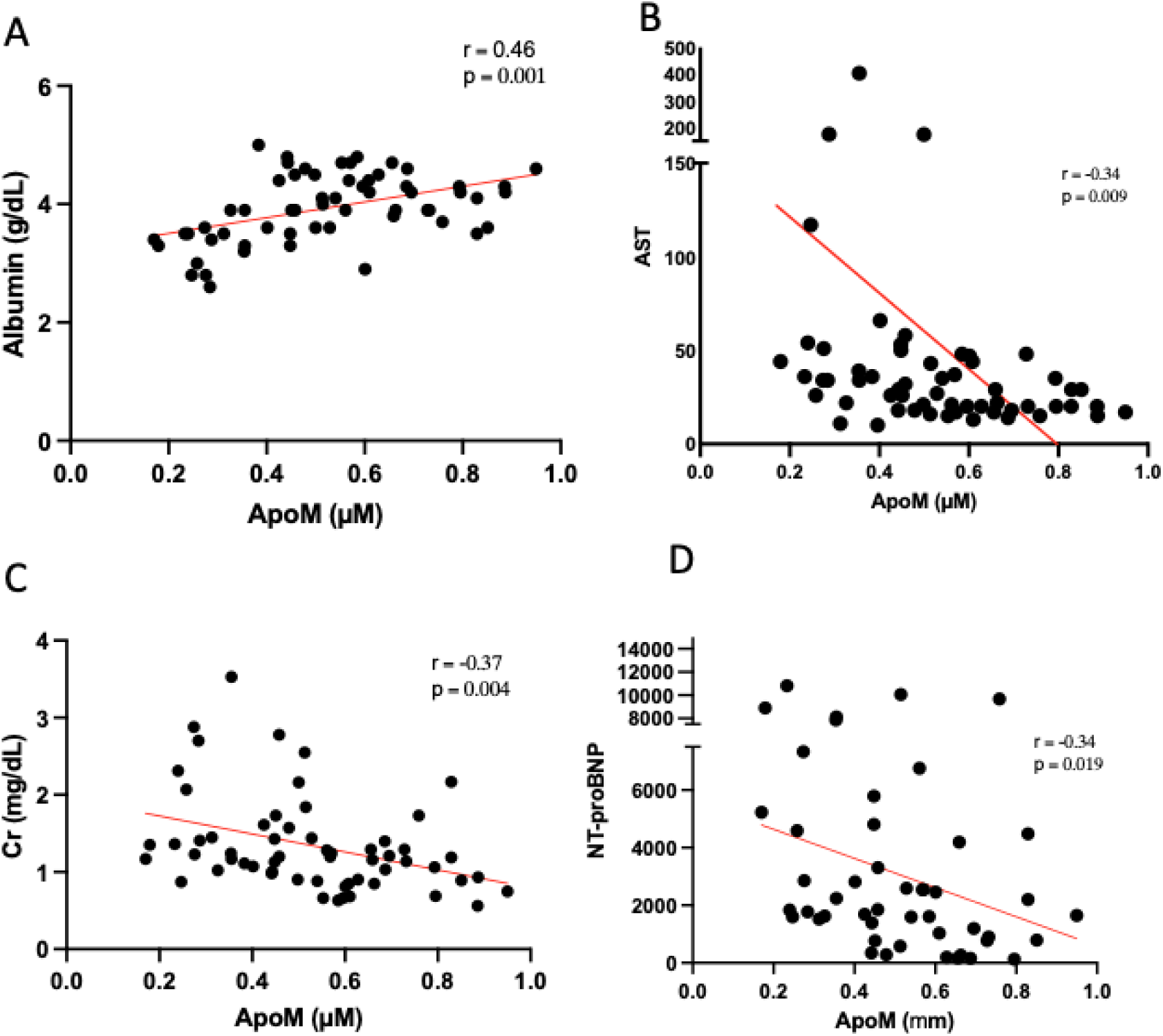
Correlation of serum ApoM levels with markers of liver and cardiac function. (A)Positive correlation with albumin and negative correlations with (B) AST, (C) creatinine, and (D) NT-proBNP. Pearson correlation coefficients (r) and p values are shown.

**Supplemental Figure 3.**
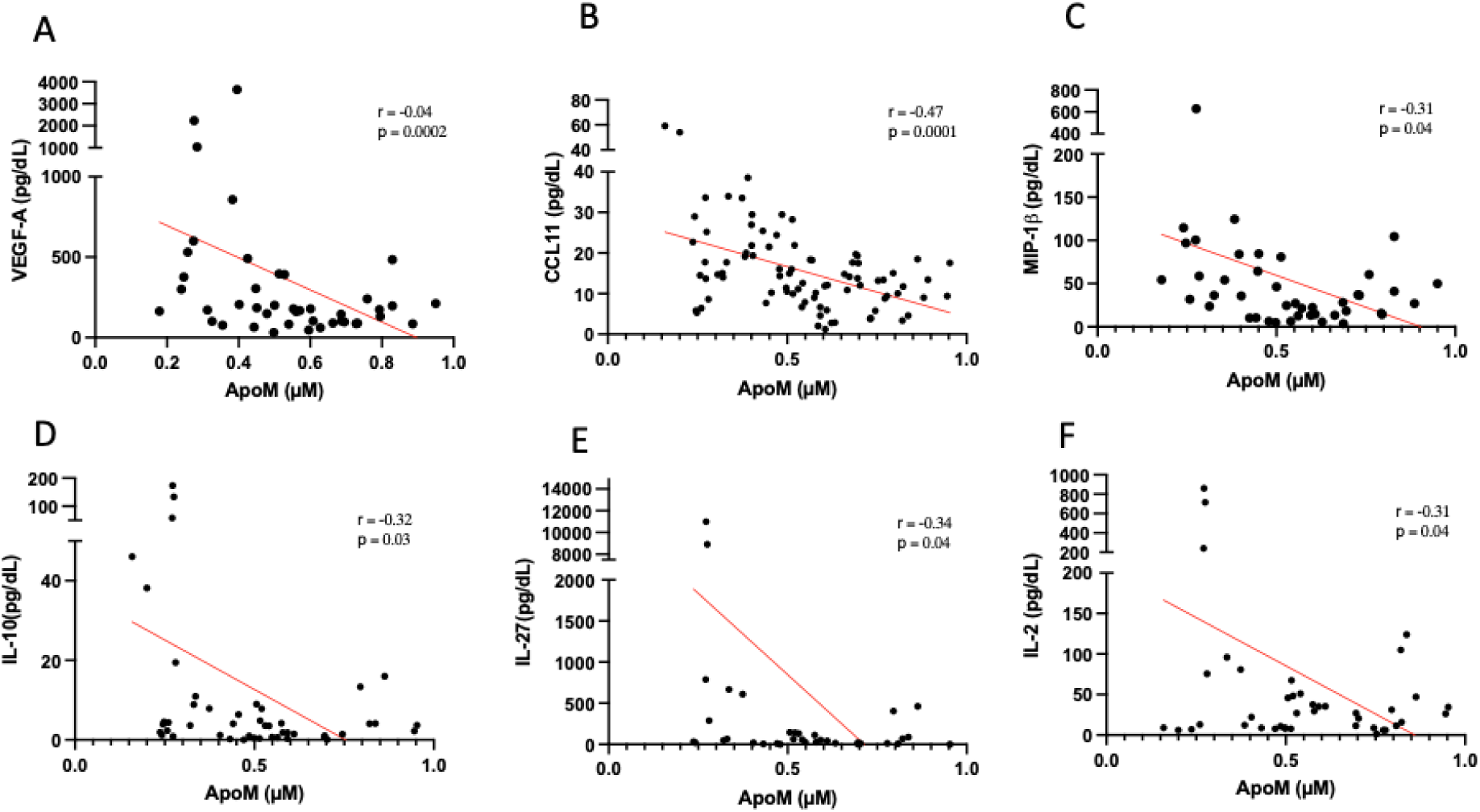
Correlation of serum ApoM levels with inflammatory markers. (A) VEGF-A, (B) CCL11, (C) MIP-1β, (D) IL-10, (E) IL-27, and (F) IL-2 show negative correlations with ApoM levels. Pearson correlation coefficients (r) and p values are indicated.

**Supplemental Figure 4.**
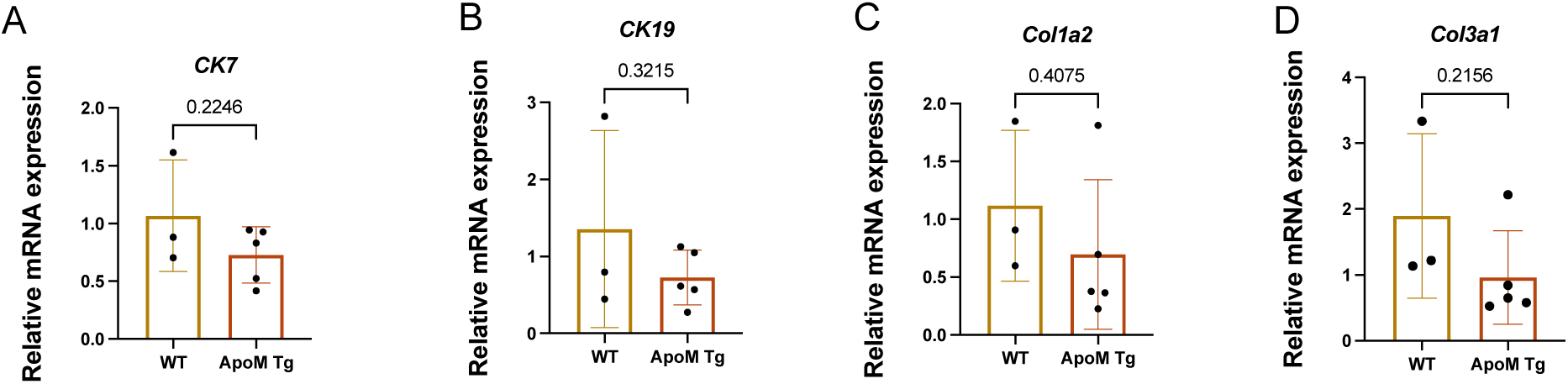
Hepatic mRNA expression of ductular reaction and fibrosis-associated genes in ApoM transgenic (ApoM TG) and control mice. (A) *CK7*, (B) *CK19*, (C) *Col3a1*, and (D) *Col1a1*. Data are shown as relative mRNA expression (mean ± SD); p values determined by unpaired t-test.

**Supplemental Figure 5.**
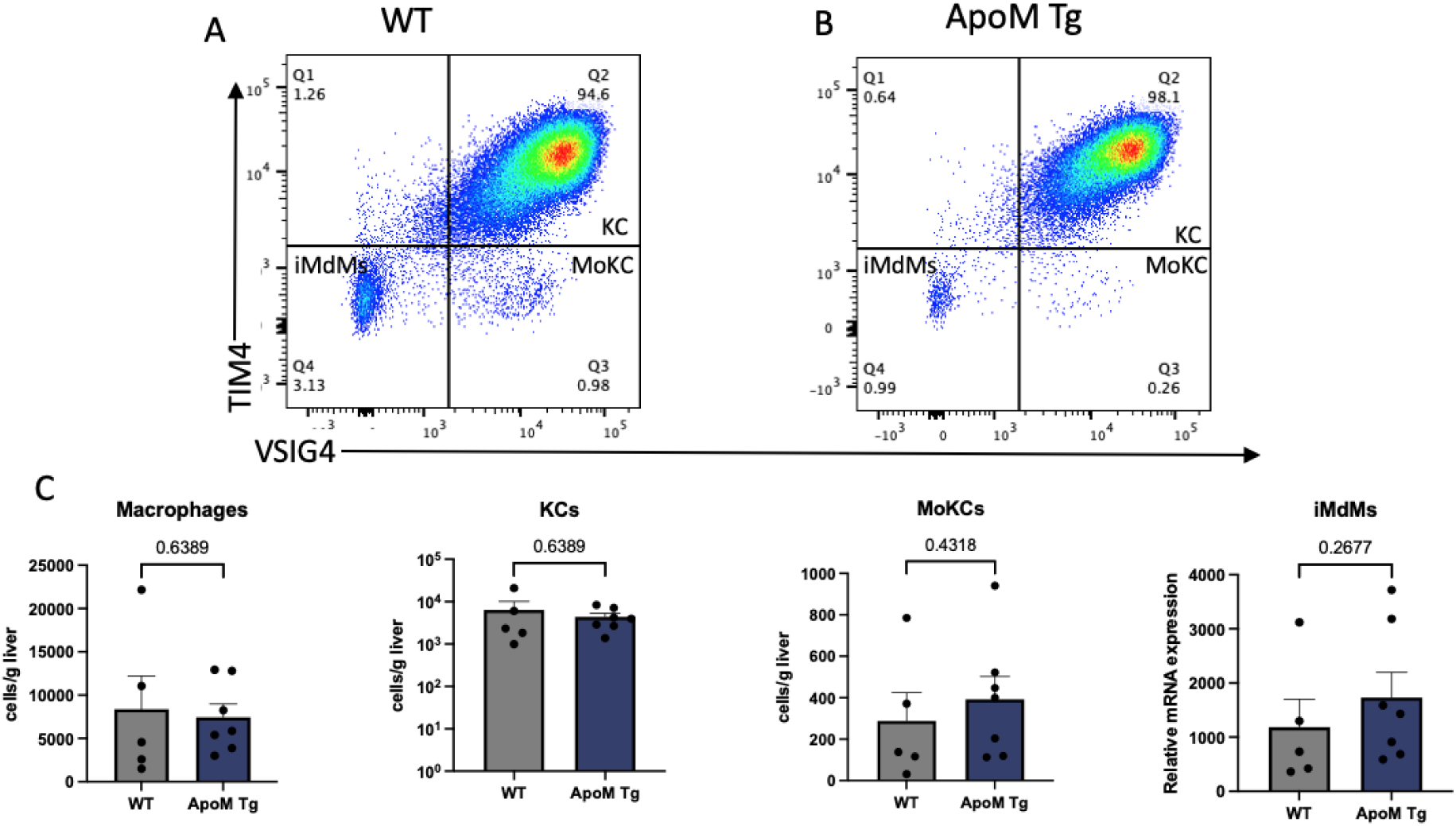
Flow cytometric analysis of hepatic macrophage populations in control and ApoM transgenic (ApoM Tg) mice. (A–B) Representative plots showing Kupffer cells (KCs), monocyte-derived Kupffer cells (MoKCs), and infiltrating monocyte-derived macrophages (iMdMs). (C) Quantification of total macrophages, KCs, MoKCs, and iMdMs per gram of liver. Data are mean ± SD; p values determined by unpaired t-test.

## Notes

### Competing Interest Statement

The authors have declared no competing interest.

### Author Declarations

This study was approved by the Washington University Institutional Review Board (No. 201903133).

## Reference

[1] Savarese G, Becher PM, Lund LH, Seferovic P, Rosano GMC, Coats AJS. Global burden of heart failure: a comprehensive and updated review of epidemiology. Cardiovasc Res 2023;118:3272–87. 10.1093/CVR/CVAC013.

[2] Bosch L, Lam CSP, Gong L, Chan SP, Sim D, Yeo D, et al. Right ventricular dysfunction in left-sided heart failure with preserved versus reduced ejection fraction. Eur J Heart Fail 2017;19:1664–71. 10.1002/EJHF.873.

[3] Ghio S, Temporelli PL, Klersy C, Simioniuc A, Girardi B, Scelsi L, et al. Prognostic relevance of a non-invasive evaluation of right ventricular function and pulmonary artery pressure in patients with chronic heart failure. Eur J Heart Fail 2013;15:408–14. 10.1093/EURJHF/HFS208.

[4] Park AC, Schilling JD. The Cardiohepatic Axis in Heart Failure. JACC Basic Transl Sci 2025;10:101312. 10.1016/J.JACBTS.2025.05.007.

[5] Laribi S, Mebazaa A. Cardiohepatic syndrome: Liver injury in decompensated heart failure. Curr Heart Fail Rep 2014;11:236–40. 10.1007/S11897-014-0206-8/FIGURES/1.

[6] Park AC, Fu CF, Parvathaneni A, Yang BQ, Chan MM, Byrnes K, et al. Biliary Metaplasia and Macrophage Activation Define the Cellular Landscape of Cardiogenic Liver Disease. JACC Basic Transl Sci 2025;10:434–54. 10.1016/J.JACBTS.2024.12.003.

[7] Yang BQ, Park AC, Liu J, Byrnes K, Javaheri A, Mann DL, et al. Distinct Inflammatory Milieu in Patients with Right Heart Failure. Circ Heart Fail 2023;16:E010478. 10.1161/CIRCHEARTFAILURE.123.010478/FORMAT/EPUB.

[8] Ren K, Tang ZL, Jiang Y, Tan YM, Yi GH. Apolipoprotein M. Clin Chim Acta 2015;446:21–9. 10.1016/J.CCA.2015.03.038.

[9] Christoffersen C, Nielsen LB. Apolipoprotein M: bridging HDL and endothelial function. Curr Opin Lipidol 2013;24:295–300. 10.1097/MOL.0B013E328361F6AD.

[10] Liu M, Seo J, Allegood J, Bi X, Zhu X, Boudyguina E, et al. Hepatic Apolipoprotein M (ApoM) Overexpression Stimulates Formation of Larger ApoM/Sphingosine 1-Phosphate-enriched Plasma High Density Lipoprotein. Journal of Biological Chemistry 2014;289:2801–14. 10.1074/JBC.M113.499913.

[11] Christoffersen C, Nielsen LB. Apolipoprotein M - a new biomarker in sepsis. Crit Care 2012;16:126-. 10.1186/CC11320/METRICS.

[12] Bisgaard LS, Christoffersen C. Apolipoprotein M/sphingosine-1-phosphate: Novel effects on lipids, inflammation and kidney biology. Curr Opin Lipidol 2019;30:212–7. 10.1097/MOL.0000000000000606.

[13] Tydén H, Lood C, Jönsen A, Gullstrand B, Kahn R, Linge P, et al. Low plasma concentrations of apolipoprotein M are associated with disease activity and endothelial dysfunction in systemic lupus erythematosus. Arthritis Res Ther 2019;21:110-. 10.1186/S13075-019-1890-2/FIGURES/4.

[14] Bosteen MH, Madsen Svarrer EM, Bisgaard LS, Martinussen T, Madsen M, Nielsen LB, et al. Effects of apolipoprotein M in uremic atherosclerosis. Atherosclerosis 2017;265:93–101. 10.1016/J.ATHEROSCLEROSIS.2017.08.005.

[15] Chirinos JA, Zhao L, Jia Y, Frej C, Adamo L, Mann D, et al. Reduced Apolipoprotein M and Adverse Outcomes Across the Spectrum of Human Heart Failure. Circulation 2020;141:1463–76. 10.1161/CIRCULATIONAHA.119.045323/SUPPL_FILE/SUPPLEMENTAL.

[16] Xu N, Hurtig M, Zhang XY, Ye Q, Nilsson-Ehle P. Transforming growth factor-beta down-regulates apolipoprotein M in HepG2 cells. Biochim Biophys Acta Mol Cell Biol Lipids 2004;1683:33–7. 10.1016/j.bbalip.2004.04.001.

[17] Ren K, Mo ZC, Liu X, Tang ZL, Jiang Y, Peng XS, et al. TGF-β Down-regulates Apolipoprotein M Expression through the TAK-1-JNK-c-Jun Pathway in HepG2 Cells. Lipids 2017;52:109–17. 10.1007/s11745-016-4227-9.

[18] Christoffersen C, Nielsen LB, Axler O, Andersson A, Johnsen AH, Dahlbäck B. Isolation and characterization of human apolipoprotein M-containing lipoproteins. J Lipid Res 2006;47:1833–43. 10.1194/jlr.M600055-JLR200.

[19] Guo Z, Valenzuela Ripoll C, Picataggi A, Rawnsley DR, Ozcan M, Chirinos JA, et al. Apolipoprotein M Attenuates Anthracycline Cardiotoxicity and Lysosomal Injury. JACC Basic Transl Sci 2023;8:340–55. 10.1016/J.JACBTS.2022.09.010.

[20] Mamazhakypov A, Veith C, Schermuly RT, Sydykov A. Surgical protocol for pulmonary artery banding in mice to generate a model of pressure-overload-induced right ventricular failure. STAR Protoc 2023;4:102660. 10.1016/J.XPRO.2023.102660.

[21] Simonetto DA, Yang H yin, Yin M, de Assuncao TM, Kwon JH, Hilscher M, et al. Chronic passive venous congestion drives hepatic fibrogenesis via sinusoidal thrombosis and mechanical forces. Hepatology 2015;61:648–59. 10.1002/HEP.27387/SUPPINFO.

[22] Velagapudi S, Rohrer L, Poti F, Feuerborn R, Perisa D, Wang D, et al. Apolipoprotein M and Sphingosine-1-Phosphate Receptor 1 Promote the Transendothelial Transport of High-Density Lipoprotein. Arterioscler Thromb Vasc Biol 2021;41:E468–79. 10.1161/ATVBAHA.121.316725/FORMAT/EPUB.

[23] Yao Mattisson I, Christoffersen C. Apolipoprotein M and its impact on endothelial dysfunction and inflammation in the cardiovascular system. Atherosclerosis 2021;334:76–84. 10.1016/J.ATHEROSCLEROSIS.2021.08.039.

[24] Ruiz M, Frej C, Holmér A, Guo LJ, Tran S, Dahlbäck B. High-density lipoprotein-associated apolipoprotein M limits endothelial inflammation by delivering sphingosine-1-phosphate to the sphingosine-1-phosphate receptor 1. Arterioscler Thromb Vasc Biol 2017;37:118– 29. 10.1161/ATVBAHA.116.308435.

[25] Borup A, Christensen PM, Nielsen LB, Christoffersen C. Apolipoprotein M in lipid metabolism and cardiometabolic diseases. Curr Opin Lipidol 2015;26:48–55. 10.1097/MOL.0000000000000142.

[26] Yao Mattisson I, Christoffersen C. Apolipoprotein M and its impact on endothelial dysfunction and inflammation in the cardiovascular system. Atherosclerosis 2021;334:76– 84. 10.1016/J.ATHEROSCLEROSIS.2021.08.039.

[27] Liu M, Seo J, Allegood J, Bi X, Zhu X, Boudyguina E, et al. Hepatic apolipoprotein M (ApoM) overexpression stimulates formation of larger ApoM/sphingosine 1-phosphate-enriched plasma high density lipoprotein. Journal of Biological Chemistry 2014;289:2801–14. 10.1074/jbc.M113.499913.

[28] Zhang H, Pluhackova K, Jiang Z, Böckmann RA. Binding Characteristics of Sphingosine-1-Phosphate to ApoM hints to Assisted Release Mechanism via the ApoM Calyx-Opening. Scientific Reports 2016 6:1 2016;6:30655-. 10.1038/srep30655.

[29] Cheng G, Zheng L. Regulation of the apolipoprotein M signaling pathway: a review. J Recept Signal Transduct Res 2022;42:285–92. 10.1080/10799893.2021.1924203.

[30] McLean P, Mathew D, Chandrasekhar S, Guynn N, Razavi AC, Quyyumi A, et al. HDL-C: Helpful, harmful, hopeful, or guilty by association. J Clin Lipidol 2025. 10.1016/J.JACL.2025.09.017.

[31] Franczyk B, Młynarska E, Rysz-Górzyńska M, Gluba-Sagr A, Rysz J, Sheth S, et al. High-density lipoproteins, Part 1. Epidemiology, antiatherogenic effects, and therapies designed to increase their serum levels. Am J Prev Cardiol 2025;23:101068. 10.1016/J.AJPC.2025.101068.

[32] Bril F, Pearce RW, Collier TS, McPhaul MJ. Differences in HDL-Bound Apolipoproteins in Patients With Advanced Liver Fibrosis Due to Nonalcoholic Fatty Liver Disease. J Clin Endocrinol Metab 2022;108:42–51. 10.1210/CLINEM/DGAC565.

[33] Murphy SP, Kakkar R, McCarthy CP, Januzzi JL. Inflammation in Heart Failure: JACC State-of-the-Art Review. J Am Coll Cardiol 2020;75:1324–40. 10.1016/J.JACC.2020.01.014/ASSET/BECA502B-CFEB-40DE-89B7-112051C32B50/ASSETS/GRAPHIC/GR3.JPG.

[34] Li T, Yang L, Zhao S, Zhang S. Correlation Between Apolipoprotein M and Inflammatory Factors in Obese Patients. Medical Science Monitor 2018;24:5698–703. 10.12659/MSM.907744.

[35] Mamazhakypov A, Veith C, Schermuly RT, Sydykov A. Surgical protocol for pulmonary artery banding in mice to generate a model of pressure-overload-induced right ventricular failure. STAR Protoc 2023;4:102660. 10.1016/J.XPRO.2023.102660.

[36] Chan MM, He L, Finck BN, Schilling JD, Daemen S. Cutting Edge: Hepatic Stellate Cells Drive the Phenotype of Monocyte-derived Macrophages to Regulate Liver Fibrosis in Metabolic Dysfunction-associated Steatohepatitis. J Immunol 2024;213:251–6. 10.4049/JIMMUNOL.2300847.

[37] X Z, Y K, Y S, S L, T S. Liver Fibrosis: Interactions Between Cells and Microenvironments. Turk J Gastroenterol 2025;36:711. 10.5152/TJG.2025.25313.

